# MULTIMODAL EEG WORLD MODEL: SELF-SUPERVISED LATENT TRANSITION LEARNING FOR WEARABLE EEG SEIZURE DETECTION

**DOI:** 10.64898/2026.07.20.26358450

**Authors:** Ziyuan Yin, Yifei Bai, Luqi Zhou, Hongyi Zhu, John Moraros, Guangliang Cheng, Shuihua Wang

**Affiliations:** Xi’an Jiaotong-Liverpool University, Suzhou, China; Apon Medical Ltd, Shanghai, China; University of Liverpool, Liverpool, UK; ShanghaiTech University, Shanghai, China

**Keywords:** Multimodal EEG, World Model, Self-Supervised Learning, Latent Space Learning, Seizure Detection

## Abstract

Existing supervised and self-supervised EEG models mainly learn within-segment discriminative or reconstructive representations, leaving transitions between adjacent EEG segments underexplored. We propose a self-supervised Multimodal EEG World Model for EEG seizure detection that predicts the latent representation of the next EEG segment from the current EEG state, conditioned on synchronized ECG, EMG, and movement signals. A learnable query-based fusion module aggregates the multimodal synchronized signals into a compact physiological condition. Pretraining uses no seizure labels; downstream detection retains only the frozen EEG encoder and a linear probe. On the SeizeIT2 dataset with a strict patient-wise split, the model achieved an AUPRC of 0.3748±0.0127, ROC-AUC of 0.8025±0.0071, and balanced accuracy of 0.7308±0.0075, outperforming official database baseline (SVM with hand-crafted features, ChronoNet), external baselines (DSAINet) and foundation models (NeuroLM). Ablations show a 14.4% relative AUPRC gain over a random-initialized encoder, supporting multimodal-conditioned latent transition learning for wearable EEG representation learning.

## 1. INTRODUCTION

### 1.1 EEG Seizure Detection

Epilepsy is a neurological disorder characterized by abnormal neuronal discharges, which may lead to involuntary muscle contractions, impaired consciousness, and an increased risk of injury and mortality. Globally, approximately 0.7% of the population is affected by epilepsy [1]. Automated seizure detection systems have the potential to improve long-term disease monitoring and enhance the quality of life of patients with epilepsy [2].

### 1.2. Limitations of Existing Seizure Detection Models

Automated seizure detection algorithms have evolved through several stages. Early approaches mainly relied on statistical or machinelearning classifiers [3, 4]. Later, deep learning models were introduced, including recurrent architectures such as ChronoNet [5]. More recently, EEG foundation models based on large-scale selfsupervised pretraining have attracted increasing attention [6].

Despite these advances, most existing EEG representation-learning approaches primarily focus on modeling information contained within individual EEG segments. Reconstruction-based methods learn representations by recovering missing information from the current observation. More recent joint-embedding predictive architectures (JEPA) avoid direct reconstruction in the raw signal space and instead perform representation prediction in a lower-dimensional latent space [7, 8].

Nevertheless, most existing JEPA-based EEG models mainly perform within-segment latent completion, where latent representations of masked regions are inferred from the remaining content of the same EEG segment [9]. They do not explicitly model the latent state transition between temporally adjacent EEG segments.

This distinction is relevant to seizure detection because seizure-related EEG activity is temporally evolving rather than static within an isolated segment. Previous studies suggested that incorporating information from temporal neighborhood improves performance [10], indicating that changes across neighboring EEG segments may contain seizure-discriminative information. This leaves an important research gap: how to explicitly learn segment-to-segment latent transition information for EEG seizure detection.

### 1.3. Contributions

We propose a self-supervised Multimodal EEG World Model for seizure detection based on Le World Model [11]. The model predicts the latent state of next-segment EEG from the latent state of current-segment EEG, conditioned on cross-attention of synchronized physiological information from ECG, EMG, and MOV. A learnable query based multimodal fusion module is introduced to aggregate heterogeneous auxiliary physiological signal into a compact condition for EEG latent state prediction.

We conduct a systematic evaluation on the SeizeIT2 dataset [12]. We retained the official SeizeIT2 training subjects (sub-001–096) and further divided the original validation subjects into validation (sub-097–102) and held-out test (sub-103–125).

It outperforms official database baseline ChronoNet by 11.1% ROC-AUC and 25.6% AUPRC; external baselines (DSAINet) by 6.3% ROC-AUC and 7.8% AUPRC; and foundation models (NeuroLM) by 14.9% ROC-AUC and 82% AUPRC. Further ablation study reveals that our proposed method achieved the highest AUPRC, ROC-AUC, and balanced accuracy, with a relative AUPRC improvement of 14.4% and ROC-AUC improvement of 2.9% over the random-initialized control encoder. Such ablation results strongly suggest the effectiveness of our proposed method.

## 2. METHOD

The overview of the proposed model’s architecture and training pipelines are shown in Fig.1.

**Fig. 1.**
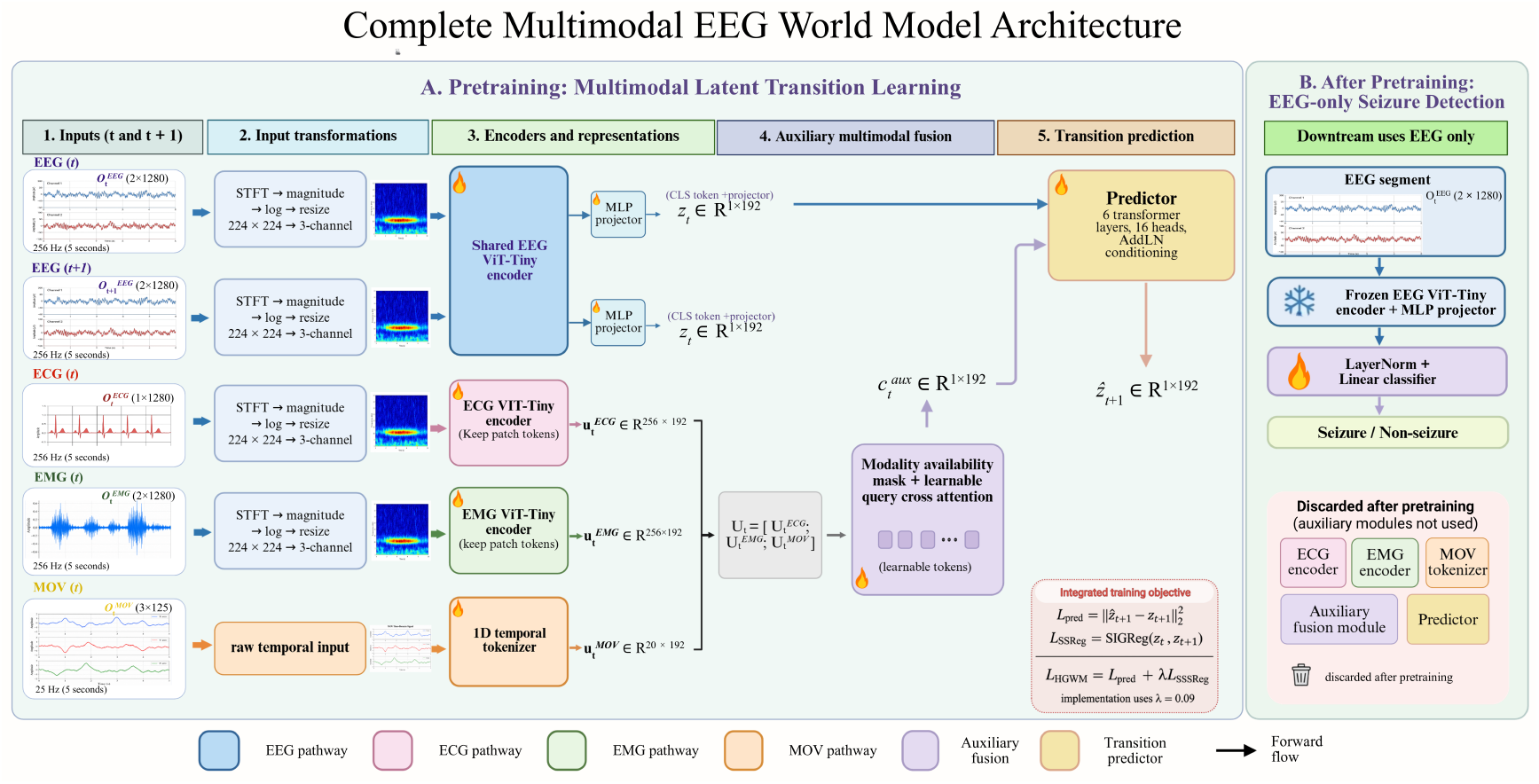
Overview of the proposed model architecture and training pipeline. Generated with BioRender.

### 2.1. Multimodal EEG World Model Architecture and Objectives

#### Architecture

Given temporally aligned multimodal recordings, each training sample is formulated as:

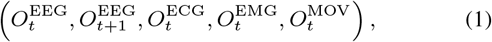

where 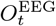 and 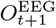 denote two consecutive EEG segments, and 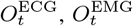, and 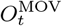 denote the auxiliary physiological signals synchronized with the current EEG segment.

For the EEG branch, each two-channel EEG segment is converted into a time-frequency representation using short-time fourier transform, followed by magnitude extraction, logarithmic scaling, resizing to 224 × 224, and conversion to a three-channel input. The current and next EEG segments are then encoded by the same EEG encoder, i.e. Vision Transformer (ViT):

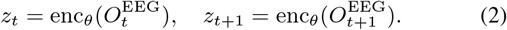

The EEG encoder is implemented as a ViT-Tiny initialized from scratch, with a patch size of 14; 12 layers; 3 attention heads, and a hidden dimension of 192. The final layer’s CLS token is passed through an MLP projection head to obtain the EEG latent representation.

The auxiliary ECG and EMG signals are also converted into time-frequency representations using short-time fourier transform and processed by two independently parameterized ViT-Tiny encoders. Instead of using the CLS token of final layer, the auxiliary encoders retain the patch-level representations:

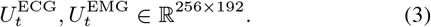

The MOV signal remains in the one-dimensional temporal domain and is processed by a temporal tokenizer consisting of two onedimensional convolutional layers followed by adaptive pooling and linear projection. This produces:

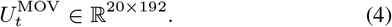

The ECG, EMG, and MOV tokens are concatenated to form the auxiliary multimodal token sequence:

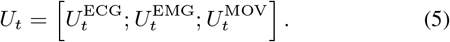

To aggregate the heterogeneous auxiliary representations, we introduce a learnable query-based physiological bottleneck containing eight learnable queries. The queries attend to the multimodal token sequence through three-head cross-attention:

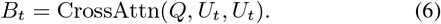

A learnable scoring function assigns an importance weight to each physiological query representation. The weighted query representations are then aggregated and projected into a single multimodal condition:

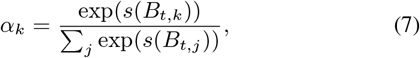

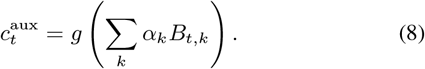

The predictor then estimates the latent representation of the next EEG segment from the current EEG latent representation and the multimodal physiological condition:

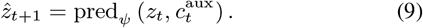

The predictor contains 6 transformer layers and 16 attention heads, and the multimodal condition is injected into the predictor through adaptive layer normalization. The EEG encoder, ECG encoder, EMG encoder, MOV tokenizer, multimodal fusion module, projection heads, and predictor are optimized jointly during pretraining. No seizure label is provided to the predictor or used as an optimization target during representation learning.

#### Prediction loss

The prediction loss is defined as the mean squared error between the predicted next-segment embedding and the encoded target embedding:

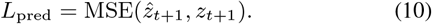

This term encourages the encoder and predictor to learn latent EEG representations that are informative for short-range temporal evolution.

#### SIGReg regularization

A prediction loss alone can lead to degenerate solutions in which the encoder maps different inputs to similar embeddings. To prevent collapse, EEG World Model applies SIGReg to the sequence of latent embeddings of the current and next segments:

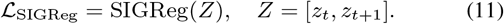

SIGReg regularizes the embedding distribution toward an isotropic gaussian distribution, promoting feature diversity in the latent space. *Total objective*.: The final objective is:

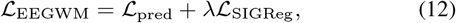

where *λ* controls the strength of SIGReg regularization.

### 2.2. Training Pipeline

The training pipeline consists of two stages. In the first stage, Multimodal EEG World Model is pretrained through multimodal-conditioned latent transition learning where predictor predicts the next-segment EEG conditioned on synchronized ECG, EMG and MOV. In the second stage, only the pretrained EEG encoder and projection head are retained and frozen for linear binary probing for seizure detection.

#### Stage 1:Pretraining

All pretraining components are optimized jointly using the EEG World Model objective Eq. (12) defined in Section 2.1. The checkpoint with the lowest validation loss is retained.

#### Stage 2:Supervised Linear Probing

This linear probing process is trained on the same training, validation set as pretraining process, but tested on a completely independent test dataset as shown in Table 1. A lightweight classifier head is trained on top of the frozen encoder. The classifier head consists of layer normalization followed by a linear projection layer. The classifier head is optimized with binary cross-entropy with logits.

**Table 1.** Patient-wise split and seizure EEG duration used in this study.

| Split | Subjects | Subject IDs | Duration (s/h) |
| --- | --- | --- | --- |
| Training | 96 | sub-001–sub-096 | 41,895 / 11.64 |
| Validation | 6 | sub-097–sub-102 | 1,298 / 0.36 |
| Test | 23 | sub-103–sub-125 | 8,382 / 2.33 |
| Total | 125 | sub-001–sub-125 | 51,575 / 14.33 |

The checkpoint with the highest AUPRC on validation set is selected for final testing.

## 3. EXPERIMENTS

### 3.1. Dataset

We adopted the SeizeIT2 dataset, a multimodal dataset recorded from patients with focal epilepsy from wearable devices [12]. SeizeIT2 contains more than 11,000 hours of data from 125 patients including EEG, ECG, EMG, and movement. The dataset contains 883 focal seizure events. The detailed patient-wise split is shown in Table 1.

All recordings were preprocessed into 5-second segments. EEG were filtered between 0.5 and 60 Hz. ECG were filtered between 0.5 and 40 Hz, while EMG were filtered between 20 and 100 Hz. EEG, ECG and EMG were also filtered with 50-Hz notch filter. MOV signals were segmented directly without filtering. EEG, ECG, and EMG were sampled at 256 Hz, MOV was sampled at 25 Hz.

### 3.2. Ablation Study

#### Multimodal EEG World Model

This is the proposed complete model. The predictor estimates the next EEG latent representation from the current EEG latent representation conditioned on the fused ECG, EMG, and MOV representations. The model is pretrained using both latent transition prediction and SIGReg:

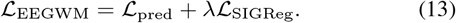

#### Label-conditioned EEG World Model

This setting replaces multimodal condition with EEG labels. The model uses the same objectives as the proposed complete model, allowing direct comparison between manually defined label conditioning and learned multimodal physiological conditioning.

#### Loss Function Only with SIGReg Control

This setting evaluates whether latent-space regularization alone is sufficient to make encoder learn useful EEG representations. In this setting, the prediction objective is removed and the EEG encoder is pretrained only using SIGReg as objective:

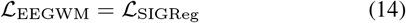

#### No-condition Control

The no-condition control removes the condition module from the predictor. The predictor therefore estimates the next EEG latent representation using only the current EEG latent representation. This setting evaluates whether conditioning module really helps the encoder to learn latent transition learning.

#### Random-Initialized Encoder Control

The random-initialized encoder control uses the same EEG encoder architecture and linear probing protocol, but the encoder is randomly initialized and frozen without representation pretraining. This setting evaluates whether the performance gain originates from the proposed pretraining procedure rather than from the encoder architecture or probing setup alone.

For clarity, the comparison of ablation settings are summarized in Table 2.

**Table 2.** Ablation and control settings used in this study.

| Setting | Condition | Objective |
| --- | --- | --- |
| Multimodal EEGWM | ECG/EMG/MOV | $\mathcal{L}_{\text{pred}} + \lambda \mathcal{L}_{\text{SIGReg}}$ |
| Label-conditioned EEGWM | EEG label | $\mathcal{L}_{\text{pred}} + \lambda \mathcal{L}_{\text{SIGReg}}$ |
| SIGReg-only | None | $\mathcal{L}_{\text{SIGReg}}$ |
| No-condition | None | $\mathcal{L}_{\text{pred}} + \lambda \mathcal{L}_{\text{SIGReg}}$ |
| Random-init. encoder | N/A | N/A |

### 3.3. Compared Baselines

External comparisons include the two official SeizeIT2 baselines, SVM with 42 handcrafted EEG features and ChronoNet [12], together with DSAINet [13] and the pretrained EEG foundation model NeuroLM [6]. We reimplemented the two official SeizeIT2 baseline methods and external models and evaluated them under our patient split and evaluation protocol.

## 4. RESULTS AND DISCUSSION

All performance are tested and reported on the independent test set as shown in Table 1. Results for models involving stochastic neuralnetwork training are reported as mean ± standard deviation across six random seeds. Deterministic baselines, including SVM and the frozen NeuroLM probing pipeline, are reported from a single run.

Ablation study results are summarized in Table 3. Comparison with external baselines is summarized in Table 4. Relative gains of our proposed model over selected methods are summarized in Table 5.

**Table 3.**
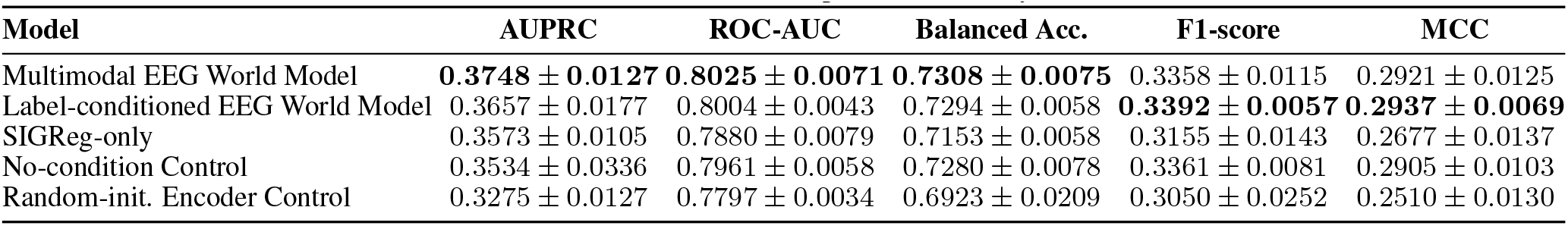
Ablation and control comparison, ranked by AUPRC.

**Table 4.** Comparison with external baselines, ranked by AUPRC.

| Model | AUPRC | ROC-AUC | Balanced Acc. | F1-score | MCC |
| --- | --- | --- | --- | --- | --- |
| Multimodal EEG World Model | **0.3748 ± 0.0127** | **0.8025 ± 0.0071** | **0.7308 ± 0.0075** | **0.3358 ± 0.0115** | 0.2921 ± 0.0125 |
| SVM | 0.3650 | 0.7494 | 0.5638 | 0.2245 | **0.2955** |
| DSAINet | 0.3476 ± 0.0107 | 0.7547 ± 0.0101 | 0.6181 ± 0.0086 | 0.3233 ± 0.0107 | 0.2760 ± 0.0176 |
| ChronoNet | 0.2984 ± 0.0191 | 0.7221 ± 0.0105 | 0.6532 ± 0.0076 | 0.2790 ± 0.0132 | 0.2044 ± 0.0143 |
| NeuroLM | 0.2059 | 0.6987 | 0.6507 | 0.2546 | 0.1820 |

**Table 5.**
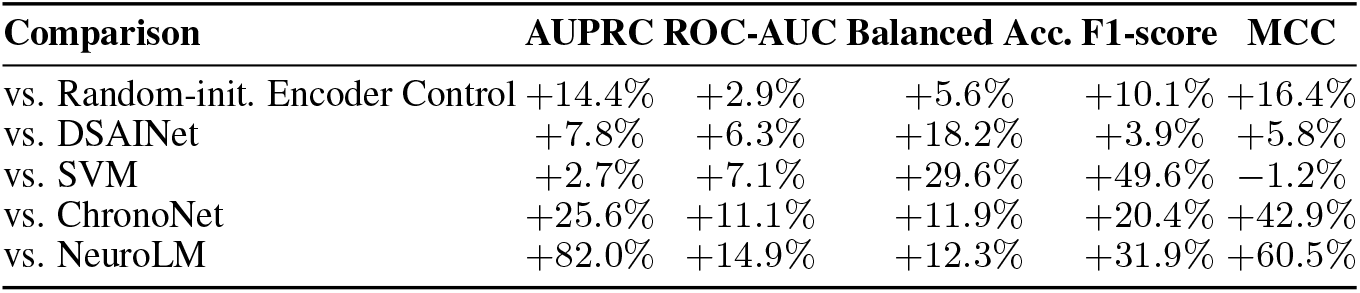
Relative gains of Multimodal EEG World Model over selected comparison methods.

| Comparison | AUPRC | ROC-AUC | Balanced Acc. | F1-score | MCC |
| --- | --- | --- | --- | --- | --- |
| vs. Random-init. Encoder Control | +14.4% | +2.9% | +5.6% | +10.1% | +16.4% |
| vs. DSAINet | +7.8% | +6.3% | +18.2% | +3.9% | +5.8% |
| vs. SVM | +2.7% | +7.1% | +29.6% | +49.6% | -1.2% |
| vs. ChronoNet | +25.6% | +11.1% | +11.9% | +20.4% | +42.9% |
| vs. NeuroLM | +82.0% | +14.9% | +12.3% | +31.9% | +60.5% |

### 4.1. Effectiveness of Multimodal Conditioning and Pretraining

The results in Table 3 show that the proposed Multimodal EEG World Model achieved the strongest AUPRC, ROC-AUC and balanced accuracy among all ablation settings. Compared with no condition control, adding synchronized multimodal physiological context improved AUPRC by approximately 6.1% in relative terms. This supports the effectiveness of our proposed multimodal condition module. This contributes to the EEG research community by pointing out a way to utilized large amount of unlabeled EEG data, i.e. pretraining the model with unlabeled EEG and synchronized auxiliary physiological signal.

The comparison with the random-initialized encoder control further demonstrates the effectiveness of our proposed whole pretraining strategy schematic including physiological contextual condition. The proposed Multimodal EEG World Model improved AUPRC by 14.4%, ROC-AUC by 2.9%, balanced accuracy by 5.6%, F1-score by 10.1%, and MCC by 16.4% relative to the random encoder control.

These results support the central motivation of this study. Seizure-related activity in EEG is not only a static pattern within an isolated 5-second segment. It may involve changes in morphology, rhythmicity, amplitude, and physiological state across neighboring segments. Multimodal latent transition pretraining encourages the EEG encoder to learn segment representations according to both temporal evolution and synchronized physiological context.

### 4.2. Role of SIGReg in Representation Learning

The loss function only with SIGReg control achieved an AUPRC of 0.3573 ± 0.0105, indicating that latent-space regularization alone can make encoder to learn useful representations EEG seizure detection.

However, SIGReg alone did not achieve the performance of the complete Multimodal EEG World Model. The proposed complete model improved over SIGReg-only setting across all five reported metrics, including relative gains of approximately 4.9% in AUPRC, 1.8% in ROC-AUC, 2.2% in balanced accuracy, 6.4% in F1-score, and 9.1% in MCC. These results suggest that SIGReg provides a useful latent-space constraint, while multimodal-conditioned next-segment latent prediction provides additional information for learning seizure-relevant EEG representations.

## 5. CONCLUSION

In this study, we proposed a self-supervised Multimodal EEG World Model for wearable seizure detection. The proposed model robustly learns to predict the next-segment EEG latent embedding from the current-segment EEG embedding conditioned on an innovative multimodal module. The synchronized multimodal physiological signals provide useful contextual information for latent EEG transition prediction. Future work will assess model’s performance on event-level metrics and diverse datasets.

## Data Availability

All data produced are available online at https://openneuro.org/datasets/ds005873/versions/1.1.0

https://openneuro.org/datasets/ds005873/versions/1.0.1/metadata

## Compliance with Ethical Standards

This study is a secondary analysis of the publicly available, deidentified SeizeIT2 dataset and involved no new participant recruitment or data collection. The original SeizeIT2 study obtained written informed consent and was approved by the UZ Leuven ethics committee (S63631); anonymization and data sharing were approved by the same committee (S67350, amendment 1) [12].

